# DNA Methylation Predicts Adverse Outcomes of Coronary Artery Disease

**DOI:** 10.1101/2023.10.07.23296703

**Authors:** Min Qin, Qili Wu, Xiaoxue Tian, Qian Zhu, Xianhong Fang, Xiaoping Chen, Chen Liu, Bin Zhang, Hanping Li, Xipei Wang, Cuiping Pan, Shilong Zhong

## Abstract

**Background:** Adverse outcomes including myocardial infarction and stroke render coronary artery disease (CAD) a leading cause of death worldwide. Biomarkers that predict such adversity enable closer medical supervision and opportunities for improved outcomes.

**Methods and results:** We present a study of genome-wide DNA methylation profiling in 933 CAD patients with up to 13 years of clinical follow-up. We discovered 115 methylation sites associated with poor prognosis and inferred that cellular senescence, inflammation, and high-density lipoprotein mediated the adversity. We built succinct prognostic models combining a few methylation sites and clinical features, which could stratify patients of different risks. Furthermore, we assessed genetic regulation of the differential methylation by interrogating QTL effects. Prognostic genes such as *FKBP5*, *UBE2E2* and *AUTS2* appeared recurrently in various analyses and were validated in patients of myocardial infarction and stroke.

**Conclusions:** Our study provides prognostic models for clinical application and revealed methylation biomarkers and mechanisms of CAD adverse outcomes.

## Introduction

Coronary artery disease (CAD) is life-threatening and represents a universal leading cause of death ^1–3^. Studies of the last century suggested a 15-year survival rate of 48-70% ^4,5^. Despite the remarkable amelioration in the recent 30 years in managing its clinical risk factors and the secondary and tertiary preventions, CAD is associated with 17.8 million annual deaths worldwide ^6,7^. Identifying patients with greater risk of poor prognosis enables closer medical supervision and therefore opportunities for better clinical outcomes. Numerous genetics-based research reported novel targets and tools for predicting adverse outcomes in CAD patients. Indeed, CAD has an estimated heritability of 0.38-0.66 for incidence ^8^ and 0.38-0.57 for mortality ^9^. However, towards which direction it progresses is multifactorial determined by the combined effects of genetic and environmental factors, therefore we reason that considering multiple layers of information, such as genetics and epigenetics, will better identify patients susceptible to poor prognostic outcomes.

DNA methylation on CpG (cytosine-phosphate-guanine) dinucleotides, a stable yet dynamic regulation mechanism reflecting both genetics and environment, enables exploring their integrated effects on diseases. Epigenome-wide association studies (EWAS) suggested DNA methylation as a feasible biomarker for CAD. Two recent large-scale EWAS surveyed multiple cohorts of various ancestries and collectively reported 85 DNA methylation sites in blood leukocytes to be associated with incident CAD or myocardial infarction (MI) ^10,11^. Comprehensive studies also report association between DNA methylation and the risk factors of CAD including aging ^12^, smoking ^13^, blood lipids ^14^, inflammation ^15^, hypertension ^16^, and diabetes mellitus (DM) ^17^. Furthermore, initial EWAS studies identified strong signals that predicted all-cause death of cardiovascular diseases ^18,19^, albeit its biological mechanisms remained to be explored. As such, DNA methylation indicates not only the risk of CAD incidence but also its progression.

In this study, we conducted a two-stage multicenter EWAS on prognosis of CAD in 933 patients, profiling blood leukocyte-derived DNA methylation by Illumina MethylationEPIC BeadChip and interrogating its association with patient outcomes in up to 13 years of follow-up. We defined the primary endpoint as all-cause death and the secondary endpoint as major adverse cardiovascular events (MACE), including death, nonfatal myocardial infarction, coronary revascularization, and stroke. From differentially methylated probes/sites (DMPs), we inferred mediating phenotypes, built risk prediction models, assessed the contribution of genetic regulation, and finally, evaluated how the genes impacted by DMPs were expressed during the adverse events. Our results show that DNA methylation of leukocytes from peripheral blood provides robust biomarkers and rich insights into the prognosis of CAD.

## Methods

### Cohort assembly and baseline information collection

This study was approved by the Medical Research Ethics Committee of Guangdong Provincial People’s Hospital (approval number: GDREC2017071H) and complied with the Declaration of Helsinki. All patients provided written informed consents.

We recruited over 5,000 CAD patients from three medical centers in two areas of China for studying the prognosis of CAD, namely Guangdong Provincial People’s Hospital, First Affiliated Hospital of Sun Yat-sen University, and Xiangya Hospital of Central South University ^20^. We selected 405 patients recruited between January 2010 to December 2017 from Guangdong Provincial People’s Hospital to form the discovery cohort, based on a nested case-control study design. 528 patients recruited from 2017 to 2018 from all three medical centers were assembled as the validation cohort. All cohort participants were identified either with a history of coronary artery bypass graft operation or newly diagnosed by coronary angiography and carotid artery ultrasonography to have ≥50% obstruction, as assessed by the luminal diameter, in minimally one main coronary artery. The inclusion criteria were: (1) aged over 30 years old, (2) no history of renal transplantation or dialysis, (3) no cirrhosis, (4) not pregnant nor breastfeeding, (5) no malignancy, (6) no history of haemodialysis; (7) no history of thyroid problems, not using antithyroid drugs nor thyroid hormone medication in the past week, and (8) completed the follow-up surveys.

The cohort participants were admitted to hospitals; after overnight fasting, their blood samples were drawn at 7AM on the second morning. Clinical laboratory tests were performed and detailed clinical surveys, including medical history, family history, smoking status, and medication intake were collected as baseline information. Echocardiography was used to determine the function and structure of the left ventricle (LV). All patients were followed up by telephone every six months by the medical staff team for inquiring about occurrences of all-cause death or major adverse cardiovascular events (MACE), with the latter defined as nonfatal myocardial infarction, coronary revascularization, stroke, and death.

### DNA extraction from blood leukocytes

Whole blood was collected in EDTA-K2 anticoagulant tubes and immediately separated into plasma and hemocyte by centrifuging at 1000 g for 10 min at 4°C. Genomic DNA was extracted from hemocyte and transferred to cryopreservation tubes, which were stored at −80°C for subsequent experiments.

### Genome-wide DNA methylation profiling and data preprocessing

DNA quality was assessed by ultraviolet spectrophotometer (Thermo Scientific, NanoDrop 2000). Briefly, about 500 ng of DNA was treated with sodium bisulfite for converting unmethylated nucleotide C to U, using the EZ DNA Methylation Kit (Zymo Research). After the conversion, methylation levels of more than 850,000 CpG sites were quantified using the Illumina Infinium MethylationEPIC BeadChip, which was run on an Illumina iScan Systems according to the manufacturer’s standard protocol. DNA methylation profiling was serviced by Genenergy Inc. The experimental operator was blind to the group information and randomly assigned the samples to different chips and plates.

Raw signal intensities of DNA methylation were stored in .idat files and imported to the R environment using the “ChAMP” package ^21,22^. Analysis was performed separately for the discovery cohort and the validation cohort. Methylation level of each probe, i.e., beta value, was defined as Meth/(Meth + Unmeth + 100), where Meth was signal intensity of the CpG site in methylated form and Unmeth was that in unmethylated form. Beta values ranged from 0 to 1, with a larger value indicating a higher level of methylation. Probes were excluded if meeting one of the following criteria: (1) with detection *P* value >= 0.01, (2) with beadcount <3 in at least 5% of samples, (3) DNA methylation occurring to non-CpG dinucleotides, (4) aligning to multiple locations ^23^, (5) located on chromosome X or Y. In total, 733,638 probes in the discovery cohort and 738,366 probes in the validation cohort were retained.

The qualified probes were normalized with the BMIQ method ^24^ to correct for signal bias caused by type-I and type-II probes on the array. Next, we used the method proposed by Houseman *et al*. ^25^ to estimate the relative proportions of blood cells, including CD8 lymphocytes, CD4 lymphocytes, natural killer cells, B cells, monocytes, and granulocytes. We also leveraged 224 positive control probes to evaluate the impact of technical confounders, which we generally referred as batch effect, on the DNA methylation values. Briefly, we computed the principal components (PCs) of these positive control probes and assessed the association between the first 20 PCs and several technical parameters, including the indices for bisulfite conversion batch, plates, sample wells and chip. Methylation residuals were then obtained via linear regression, with the beta value of each probe as independent variable, and age, sex, smoking status, estimated white-blood-cell proportions, and the top 10 PCs of positive control probes as dependent variables.

### Epigenome-wide association analysis

Cox regression-based survival analysis was employed to explore the association between each methylation residual and the trait, i.e., all-cause death or MACE. We performed such EWAS separately for the discovery and the validation cohorts. In each EWAS, we adjusted for age, sex, smoking status, percutaneous coronary intervention (PCI), arrhythmia, heart failure, hypertension, hyperlipidemia, and medication intake including β-receptor blocker (BB), angiotensin converting enzyme inhibitors (ACEI), calcium channel blocker (CCB), proton pump inhibitor (PPI), clopidogrel, and aspirin. A strict epigenome-wide significance threshold by Bonferroni correction was set as *P* < 6.83E-08 and a moderate threshold by Benjamini & Hochberg correction was set as *FDR* <0.05. The differentially methylated site was considered validated when the association showed a consistent direction of effect in both cohorts, with *FDR* <0.05 in the discovery cohort and *P* < 0.05 in the validation cohort.

### Characterizing genomic locations of DMPs

Genomic locations of DMPs were annotated by Annovar ^26^. Overlap with regulatory elements were computed based on ENCODE Encyclopedia version 5 (ENCODE5) cCRE catalog ^27^, including insulators, promoters, distal enhancers, and proximal enhancers. Enrichment against tissue- and cell type-specific regulatory elements was performed based on histone modification chromatin immunoprecipitation peaks (ChIP) (H3K4me1, H3K4me3, H3K27me3, H3K36me3, H3K9me3, and H3K27ac marks) and regions of 15 chromatin states across 299 cell types and tissues from Roadmap Epigenomics ^28,29^ in eFORGE v2.0 (https://eforge.altiusinstitute.org/).

### Target gene predictions

We predicted the target genes impacted by the DMPs by two methods. For one, we used the annotation file provided by Illumina, which assigned each CpG site to its nearest gene. For the other, we leveraged the activity-by-contact (ABC) model developed by Nasser et al. ^30,31^, which identified active enhancers in a particular cell type and predicted their target genes based on chromatin states and three-dimensional contacts. To identify the ABC enhancers that overlap with DMPs, we adopted the GWAS annotation approach by Zhang K et al. ^32^ by adding ±2500 bp to the genomic location of DMPs and overlapped them with the ABC enhancers of 131 human cell types. We adopted the original ABC score thresholds, namely ≥0.015 for distal element-gene connections and ≥0.1 for proximal promoter-gene connections, to define DMP – enhancer – target gene connections.

### Construction of prognostic models for death and MACE

Risk prediction models were constructed based on the discovery cohort and tested in the validation cohort. For building the methylation model of death, 21 DMPs passing the Bonferroni-corrected epigenome-wide significance threshold were pruned by a random forest approach (feature pruning) and those retained were fit by the multivariate Cox regression to derive the final model (weight tuning). For feature pruning, the parameters *m_try_* and *n_tree_* in the random forest models were tuned using the out of bag error for deriving a minimal overall misclassification rate. *m_try_* refers to the number of variables tried at each split and *n_tree_* refers to the number of trees to be grown in a forest. The top 10 DMPs with the largest variable importance measure (VIM), which denoted the contribution of each input feature to the model, were retained. For the risk model of MACE, all eight DMPs passing the Bonferroni-corrected epigenome-wide significance threshold were retained. In weight tuning, the retained DMPs were fit by multivariate Cox regression in the R package “survival”. For deriving robust AUC (The area under the receiver operating characteristic (ROC) curve) values, we adopted a process of 80:20 data split and 1,000 times cross validation. The final model was obtained by combining all patients in the discovery cohort.

We applied two types of models to the multi-center validation cohort for testing their performances, with one combining CpG sites, sex, and age, and the other combining CpG sites, sex, age, HDLC, Fibrinogen, and LVEF. Prediction risk scores for five-year survival were computed, and Wilcoxon test was used to assess whether the scores between the two groups of patients, i.e., with and without the events of death or MACE, were significantly different. The sensitivity and specificity of these two models were computed using the ConfusionMatrix function from the R package “caret”.

Several clinical features were also assessed for their capability in predicting CAD prognostic outcomes, alone or in combination with other clinical features and the selected methylation sites, using the R package “survival”.

### Pleiotropic association analysis of DMPs and eQTL

Summary-data based Mendelian Randomization (SMR) analysis ^33^ and the Heterogeneity in Dependent Instruments (HEIDI) test were employed to identify pleiotropic relationships between the DMPs and gene expression (https://yanglab.westlake.edu.cn/software/smr/#Download). GWAS summary statistics for DNA methylation in Asian populations, and therefore information of methylation quantitative trait loci (meQTL), were obtained from Peng et al ^34^. The association strength *beta* theoretically ranges between -1 and 1 for maximally negative to maximally positive associations. The information of *cis*-eQTL were downloaded from eQTLGen (https://www.eqtlgen.org/). Allele frequencies were obtained by referring to the East Asians in the 1000 Genomes Project reference panel (phase3, version5) ^35^. 69 DMPs with at least one *cis*-meQTL (*P* <1.0E-08) were selected for computing causal relationships with *cis*-regulated gene expression. The CpG-gene expression associations with a Bonferroni-corrected *P* value (*P* < 0.05/69 = 7.25E-04) were further selected for the HEIDI test (*P*_HEIDI_ >0.05) to distinguish pleiotropy from linkage.

### Genome-wide association study of death and MACE

We performed genome-wide association studies (GWAS) on death and MACE in 1,551 CAD patients recruited from Guangdong Provincial People’s Hospital, Xiangya Hospital of Central South University, and the First Affiliated Hospital of Sun Yat-sen University. These patients were genotyped by Illumina Infinium GSA-24 v1.0 bead chip on 700,078 single-nucleotide genomic positions, which, after genotype imputation against the East Asian populations in the 1000 Genomes Project, generated 3,435,397 high-quality single nucleotide variants (SNVs). Details about cohort enrollment, baseline characteristics, data quality control, and genotype imputation were described previously ^20^. Logistic regression was employed for the GWAS via the PLINK software (version 2.0). The first 10 principal components, sex, age, smoking status, percutaneous coronary intervention (PCI), arrhythmia, heart failure, hypertension, hyperlipidemia, and medication intake including β-receptor blocker (BB), angiotensin converting enzyme inhibitors (ACEI), calcium channel blocker (CCB), proton pump inhibitor (PPI), clopidogrel, and aspirin were included as covariates.

### Differential gene expression analysis in MI and stroke

We obtained from Kuppe et al ^36^ single-nucleus RNA sequencing data from 19 patients with acute MI and four non-transplanted heart donors as controls. A total of 191,795 nuclei from 31 tissue samples, including ten major cardiac cell types, were obtained. We performed differential gene expression analysis between the MI patients and controls, as well as among three tissue zones, namely myogenic, ischemic, and fibrotic zones. We also assessed differences between groups by cell types. Wilcoxon tests implemented in the FindMarkers function of the R package “Seurat” were used^37^. Genes passing the Bonferroni-corrected *P* value of 0.05 were considered differentially expressed. Time-series expression analysis based on Fuzzy C-means clustering was used to demonstrate the relative expression changes of prognostic genes.

We obtained bulk RNA sequencing of peripheral blood from patients of MI ^38^ and patients of ischemic stroke ^39^. Differential gene expression analysis was performed between patients and controls using the R package “limma” ^40^.

### Statistical tests

Baseline demographic and clinical characteristics were presented as mean ± standard deviation for continuous variables and counts (%) for categorical variables. Cox regression-based survival analysis was employed for assessing association between the features and outcomes. Linear regression was used to explore the relationships between DMPs and six inflammatory markers, four blood lipids and two left ventricular indices. Enrichment analysis of biological pathways and traits in GWAS Catalogue database were carried out by R package “enrichR” and terms with a *P* value smaller than 0.05 was considered as significant. Unless stated, *P* values derived from multiple tests were corrected by methods of FDR or Bonferroni correction. Wilcoxon test was used to assess if the difference of continuous variables between two groups were statistically significance. For counts, chi-square tests were used.

## Results

### Baseline characteristics of the study population

We adopted a two-stage multicenter design for studying DNA methylation related to CAD prognosis (*Figure 1a*). We assembled over 5,000 CAD patients from a large medical center in China and based on the nested case-control study design, selected 405 patients to form the discovery cohort. As such, a total of 217 deaths and 247 MACE events were recorded in up to 13 years of follow-up, while 158 patients experienced no adversity. For the validation cohort, we chose a forward study design and enrolled 528 CAD patients from three medical centers in China. We followed the patients in the validation cohort for about 3 years and observed 25 deaths and 41 MACE events.

**Figure 1.**
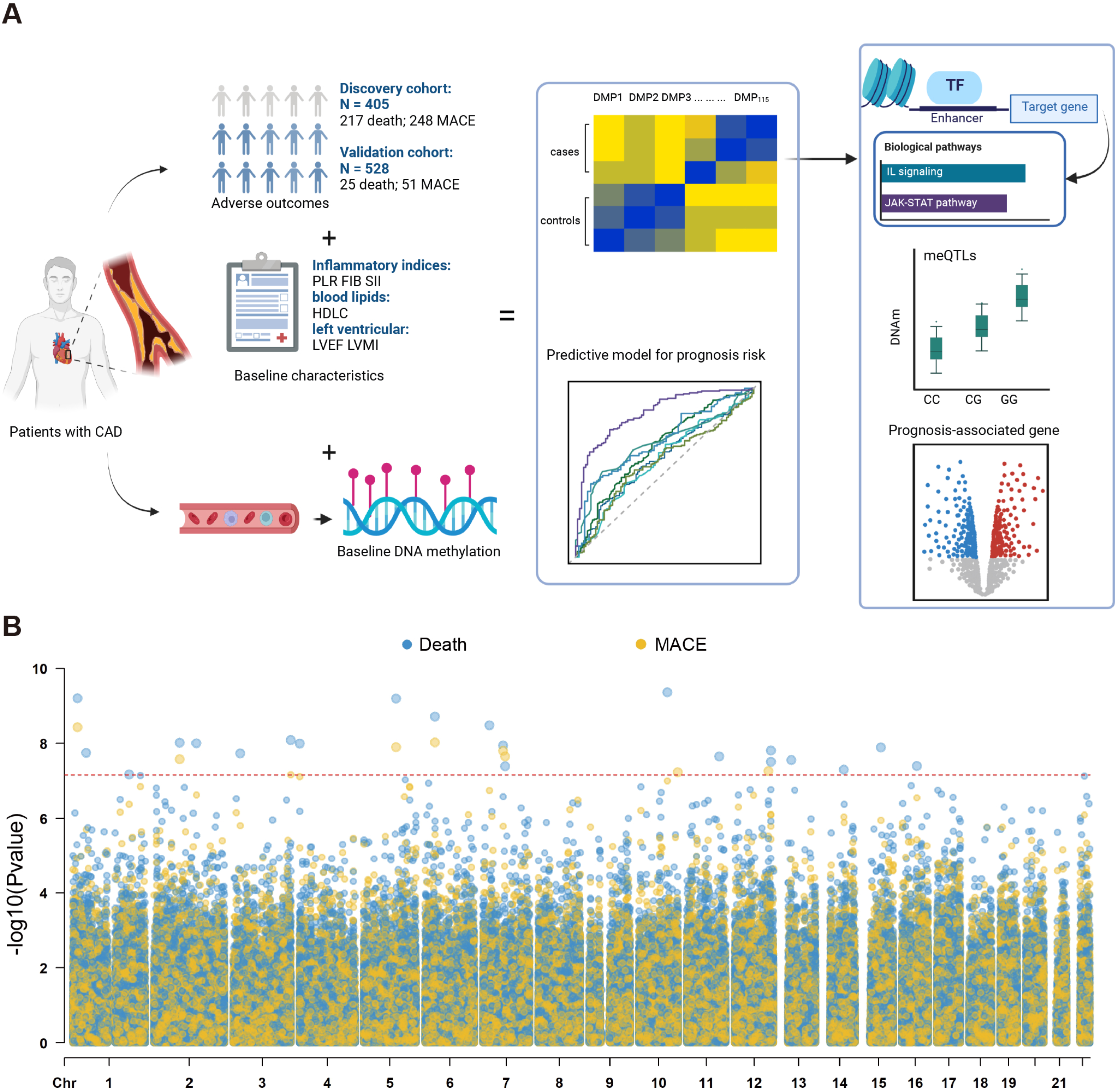
Epigenome-wide association studies on DNA methylation and CAD adverse outcomes. **(a)** Overall study design. Patients in the discovery cohort were enrolled from one medical center, whereas patients in the validation cohort were recruited from three medical centers in the China. Baseline characteristics were collected during the enrollment. DNA methylation of peripheral blood leukocytes was measured by Illumina MethylationEPIC BeadChip on ∼ 850,0000 sites. Differentially methylated sites associated with death or MACE were identified, prognostic risk models were built, and biological mechanisms were inferred. This graph was created via https://www.biorender.com/. **(b)** EWAS results of death and MACE based on the discovery cohort were presented in Manhattan plot. Red dash line marks the Bonferroni corrected *P* value threshold.

The baseline characteristics of the patients at the time of enrollment were expounded in Supplementary data online, *Table S1* and *Figure S1*. Consistent with epidemiological observations, our CAD patients were mainly men aged over 60 years (73%). We documented their basic demographic information, medical history, biomedical measurements after overnight fasting, and medication intake during the ascertained periods. The measurements were comparable between the two cohorts (*P* > 0.05), except for a few, most of which were included as covariates in our statistical tests. Overall, the two cohorts displayed similar patient characteristics. The follow-up times and event times are displayed in Supplementary data online, *Figure S1*.

### EWAS identified differentially methylated CpGs for death and MACE

We extracted DNA from leukocytes in peripheral blood and performed genome-wide methylation profiling via the Illumina Infinium MethylationEPIC BeadChip. After stringent quality control (see Supplementary data online, *Figure S2*), we obtained 733,737 and 738,021 high-quality CpG probes from the discovery and the validation cohorts, respectively.

Next, we employed Cox regression for EWAS on the high-quality DNA methylation sites against the survival of all-cause death or MACE (see Supplementary data online, *Figure S3a-S3b* and *Table S2-S4*). In the discovery cohort, after correcting for sex, age, smoking status, percutaneous coronary intervention, heart failure, hypertension, arrhythmia, hyperlipidemia, and medications, a total of 554 differentially methylated CpGs (DMPs) passed the moderate significance threshold (FDR adjusted *P*, i.e., *Q* < 0.05, *Figure 1b*). In the validation cohort, 105 of the 554 DMPs were replicated, defined by *P* < 0.05 and consistent direction of effect. Remarkably, 21 DMPs in the discovery cohort remained significant after the epigenome-wide Bonferroni correction (*P* < 6.83E-08), and six of them were replicated in the validation cohort (*P* < 0.05). Similarly, we performed EWAS on the secondary endpoint, MACE. 30 out of the 95 DMPs were validated, among which five passed the Bonferroni-corrected threshold.

Overall, 115 DMPs were detected in both cohorts as associated with adverse outcomes of CAD, with most showing increased methylation (*Figure 2a*, Supplementary data online, *Figure S3* and *Table S5*). Notably, 60% DMPs were reported in EWAS Catalog ^41^ and EWAS Atlas ^42^ to connect to a variety of traits and diseases, including Crohn’s disease and inflammatory bowel disease (25 DMPs), smoking (16 DMPs), aging (8 DMPs), and C-reactive proteins (6 DMPs) ^15^. Associations with body mass index ^17,43^, atherosclerotic plaque ^44^, and death risk ^45^ were also observed.

**Figure 2.**
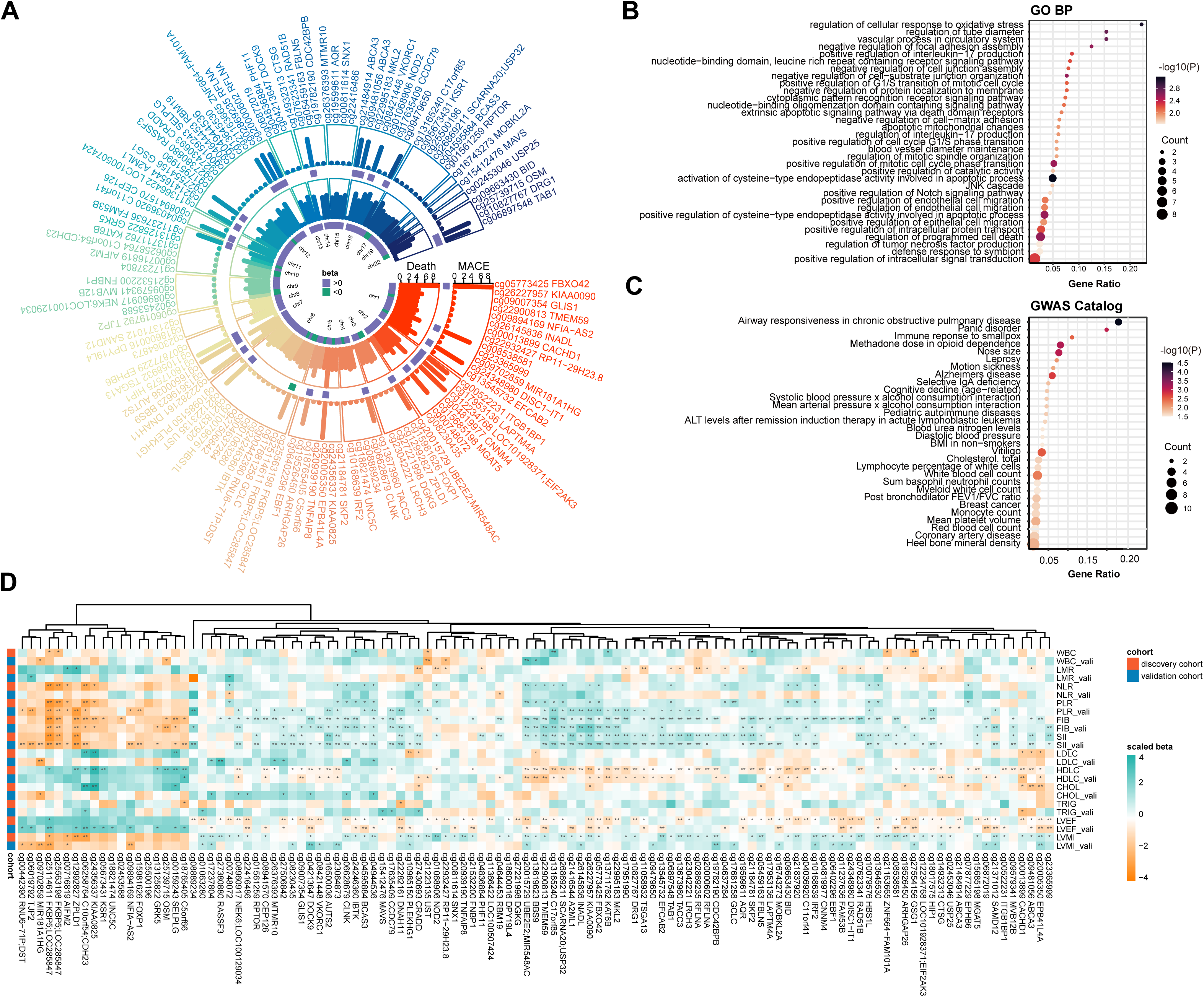
Epigenome-wide association studies of DNA methylation reveals differentially methylated CpGs associated with CAD adverse outcomes. **(a)** Genomic distribution of the 115 DMPs consistently associated with death and MACE in both the discovery and the validation cohorts. Bars represent *P* values from the discovery cohort. On the bottom of each circle, hypermethylated sites are marked in purple and hypomethylated sites in green. **(b)** Pathway enrichment of the genes connected to DMPs against the KEGG database. **(c)** Diseases or traits enriched among the genes connected to DMPs against the GWASCatalog database. **(d)** Association of the DMPs with three categories of clinical measurements: inflammation indices, lipids, and heart functions. Measurements were recorded in both the discovery (upper) and the validation (lower) cohort. WBC: whole plasma cell count, LMR: lymphocyte-monocyte ratio, NLR: neutrophil-lymphocyte ratio, PLR: platelet-lymphocyte ratio, FIB: fibrinogen, SII: systemic immune-inflammation index, LDLC: low-density lipoprotein cholesterol, HDLC: high-density lipoprotein cholesterol, CHOL: total cholesterol, TRIG: triglycerides, LVEF: left ventricular ejection fraction, LVMI: left ventricular mass index.

These 115 DMPs tend to reside in non-coding regions, particularly distal enhancers (see Supplementary data online, *Figure S4a-S4b*). Overlap with histone modification ChIP peaks and the 15 chromatin states from Roadmap (**Methods**) revealed that these DMPs were strongly enriched in enhancers specific to blood monocytes, adipocytes, myoepithelial cells, and fibroblasts. Furthermore, the left ventricle and right atrium also appeared on the top of the list (*Q* < 0.05, see Supplementary data online, *Figure S4c-S4d*). These results suggest that the DMPs were prone to occur in regions characteristic of heart traits. Note that our DNA methylation were derived from blood leukocytes; however, the DMPs tend to locate in enhancers characteristic of not only blood but also tissues and cells of non-blood origin yet known to play critical roles in CAD. Given that both DNA methylation and RNA transcription in blood were distinct from solid organs ^46^, our observation suggested that methylation in leukocytes indeed carried pathophysiological features and therefore suitable to serve as biomarkers

A main function of methylation is to regulate gene expression. By annotating DMPs to the closest genes, we discovered 100 prognostic genes, which were highly enriched for apoptotic pathways, stress response, inflammation response, and vascular processes (*Figure 2b*). Furthermore, these genes were enriched for CAD and immune traits as recorded in the GWAS Catalog (*Figure 2c*).

### Pathways and mediating phenotypes of DMPs

We also interrogated target genes by querying the connections of enhancers to target genes via the Activity-by-Contact (ABC) model, which leveraged chromatin states and three-dimensional contacts (**Methods**). For linking DMPs to enhancers, we adopted a liberal approach in Boix et al. (**Methods**) for connecting GWAS loci to enhancers, by which we extended the DMPs by adding 2500 bp to both flanking regions. Next, we assessed if the 5 Kb regions overlapped with any enhancers, which in turn were connected to genes via the ABC model. In this way, 83 of the 115 DMPs (72.1%) were connected to 806 genes (see Supplementary data online, *Table S6*). Consistent with the closest gene annotation, these genes were enriched for inflammation and cellular senescence (*P* < 0.05). Notably, three methylation sites, cg25114611, cg25500196, and cg25563198, were each predicted to interact with > 50 genes. These big gene clusters were strongly enriched in stress-induced senescence and inflammation response (*Q* < 0.05) (see Supplementary data online, *Figure S5*). Furthermore, all three DMPs were located on super-enhancers active in CAD relevant tissues, such as blood, lymphoid, adipose tissue, heart ventricle, and aorta ^47^.

Theoretically, the ABC target genes entail cell type-specific functions, due to enhancers’ selective activity in different cells. Among the blood cells, the target genes displayed an enrichment for JAK-STAT and interleukin (IL) pathways (IL-2, IL-7, IL-9, and IL-15) in monocytes and T helper cells, whereas an enrichment for IL-17 pathway was found active in more diverse cell types, including monocytes, macrophages, CD4+ T helper cells, and CD19+ B cells (see Supplementary data online, *Table S7*). We also extended the analysis to all 131 human cell types and tissues in the ABC model and found these immune pathways active in 24% of the tissues and cell, including those closely related to CAD, e.g., coronary artery, adipose, liver, epithelium, and T cells (see Supplementary data online, *Figure S6*), suggesting the adverse outcomes involved robust immune response pathways. There also appeared to be a marginal enrichment of lipid response in T cells and mitochondrial processes in monocytes and dendritic cells (*P* < 0.05).

We verified the connection of the DMPs to inflammation and lipids in clinical measurements (see *Figure 2d* and Supplementary data online, *Table S8*). In both the discovery and the validation cohorts and out of the 115 DMPs, 43 DMPs displayed association with *systemic immune-inflammation index* (SII) as defined by neutrophil count × platelet count / lymphocyte count ^48^, 37 DMPs associated with a chronic low-grade inflammation index *fibrinogen* ^49^, and 15 DMPs associated with a marker for acute inflammation prothrombotic status *platelet-lymphocyte ratio* (PLR) ^50^. Several other inflammation markers were also investigated; however, the associations were comparably weaker and non-consistent. For lipids, a strong connection of 29 DMPs to high-density lipoprotein cholesterol (HDLC) was observed. Interestingly, few connections were found for low-density lipoprotein cholesterol (LDLC), total cholesterol, or triglycerides (TG). We also examined left ventricular ejection fraction (LVEF) and left ventricular mass index (LVMI), as low LVEF and high LVMI were indicative of LV remodeling and therefore increased risk of death or MACE in CAD. 25 and 15 DMPs were respectively associated with LVEF and LVMI, and reassuringly, most of these associations displayed opposite directions for both traits.

### Prognostic models for death and MACE in CAD

Risk prediction models help to identify CAD patients with greater risk of developing adverse outcomes. We constructed models based on DMPs that displayed epigenome-wide significance of association in the discovery cohort (n=405). We leveraged a random forest approach to select DMPs that contributed the most to the classification accuracy, and derived precise weights by the multivariable Cox regression-based survival analysis (**Methods**, see Supplementary data online, *Figure S7*). As such, we constructed prognostic models for death with 10 CpGs and for MACE with 8 CpGs, which we termed the CG prognostic models (*Table 1*). This model for death achieved an area under the curve (AUC) of 0.70 in the discovery cohort, close to or better than using traditional risk factors including sex (AUC = 0.52), chronological age (AUC = 0.72) and their combination (AUC = 0.72). Combining sex, chronological age, and the 10 CpGs improved the prediction power (AUC = 0.80) (*Figure 3a*). We also built prognostic models based on the mediating phenotypes. Although not all the clinical features were equally powerful in predicting the adverse outcomes (see Supplementary data online, *Figure S8a-S8b*), we found that the ensemble model combining the 10 DMPs, sex, age, fibrinogen, HDLC, and LVEF achieved an AUC of 0.83 (*Figure 3b*). When applying the models to the validation cohort (n=528), which was assembled from three medical centers and independent of the discovery cohort in which the models were built, we observed about 11% drop in both sensitivity and specificity. Still, the ensemble model could well stratify CAD patients of different risks of death in five years (P < 0.0001 (see *Figure 3c-3d* and Supplementary data online, *Table S9*).

**Figure 3.**
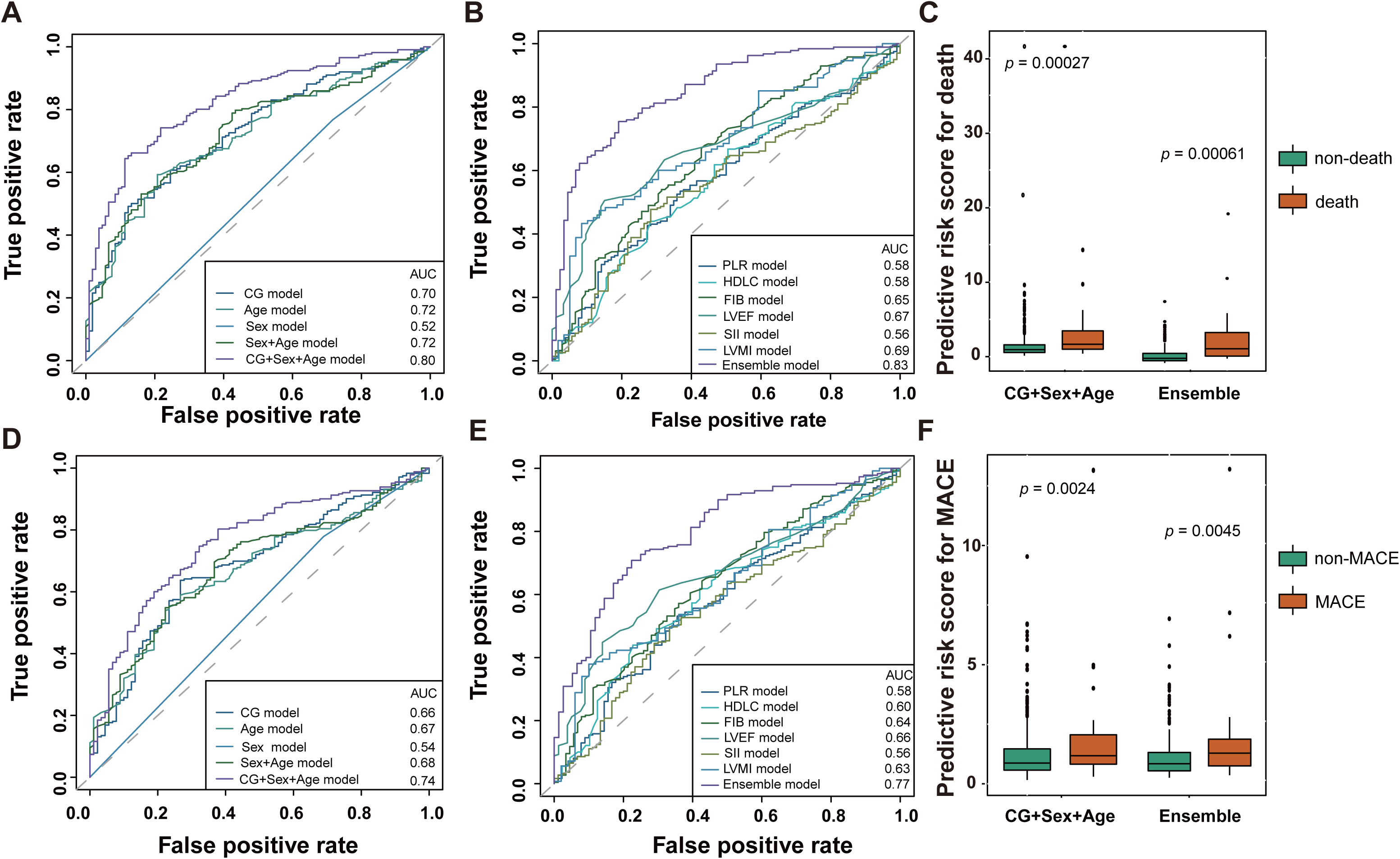
Prognostic models for CAD adverse outcomes. **(a-b)** ROC curves of the prognostic models of death constructed from the discovery cohort. The CG model consists of 10 CpG sites. The Ensemble model is composed of CG + Sex + Age + FIB + HDLC + LVEF. **(c)** Predicted risk of death in the validation cohort, by applying the model of CG + Sex + Age. **(d**) Predicted risk of death in the validation cohort, by applying the Ensemble model. **(e-f)** ROC curves of the prognostic models of MACE constructed from the discovery cohort. Features are the same as those for the model of death, except that the CG model consists of 8 CpG sites. **(g)** Predicted risk scores of MACE in the validation cohort, by applying the model of CG + Sex + Age. **(h)** Predicted risk of MACE in the validation cohort, by applying the Ensemble model. PLR: platelet-lymphocyte ratio, FIB: fibrinogen, SII: systemic immune-inflammation index, HDLC: high-density lipoprotein cholesterol, LVEF: left ventricular ejection fraction, LVMI: left ventricular mass index.

Similarly, the CG prognostic model for MACE performed the best when combining the 8 CpG sites with sex, age, fibrinogen, HDLC, and LVEF (AUC = 0.77) (*Figure 3e-3f*) and achieved a good patient stratification of five-year risk in the validation cohort (see *Figure 3g-3h* and Supplementary data online, *Table S10*).

Age is known as a strong risk factor for CVD. Observing the chronological age predicted closely to the CG models, we explored the performance of several DNA methylation clocks, including GrimAge ^51^, PhenoAge ^52^, Hannum Clock ^53^, and Horvath Clock ^54^. DNA methylation clocks have been shown to better represent one’s aging status. Indeed, most clock models achieved better prediction than the chronological age models for death and MACE (see Supplementary data online, *Figure S8c-S8d*) and performed equally well or even better than the CG prognostic models. As these clocks comprised dozens to hundreds of CpG sites, one to two orders of magnitude more than the maximal 10 CpG sites in our models, our CG prognostic models are more succinct and specific.

Note that among our CAD patients with adverse outcomes, a vast majority had the events occurred within the first 5 years of the ascertained period, therefore our models mostly captured the risk of adversity in a relative near term.

### Contribution of genetic regulation to CAD prognosis

Methylation can be regulated genetically by quantitative trait loci (meQTL), thus providing a tool for investigating how genetics influences CAD prognosis. We queried the 115 DMPs against a meQTL dataset derived from 3,523 East Asians (**Methods**). Compared with all CpG sites on the Infinium methylationEPIC beadchip, the DMPs were enriched for both *cis*-meQTL, defined as within 1 Mb flanking regions (Hypergeometric *P* = 2.12E-08), and *trans*-meQTL, defined as >5 Mb or SNP-CpG pair located on different chromosomes (Hypergeometric *P* = 3.55E-26). Briefly, 70 DMPs were subjected to the regulation of 14,374 unique *cis*-meQTLs and 52 DMPs to 1,612 unique *trans*-meQTLs, with weak associations in most cases. Compared with *trans*, DMPs in *cis* relations tended to have more meQTLs. Altogether 92 of the 115 DMPs (80%) were regulated genetically (*Figure 4a-4d*).

**Figure 4.**
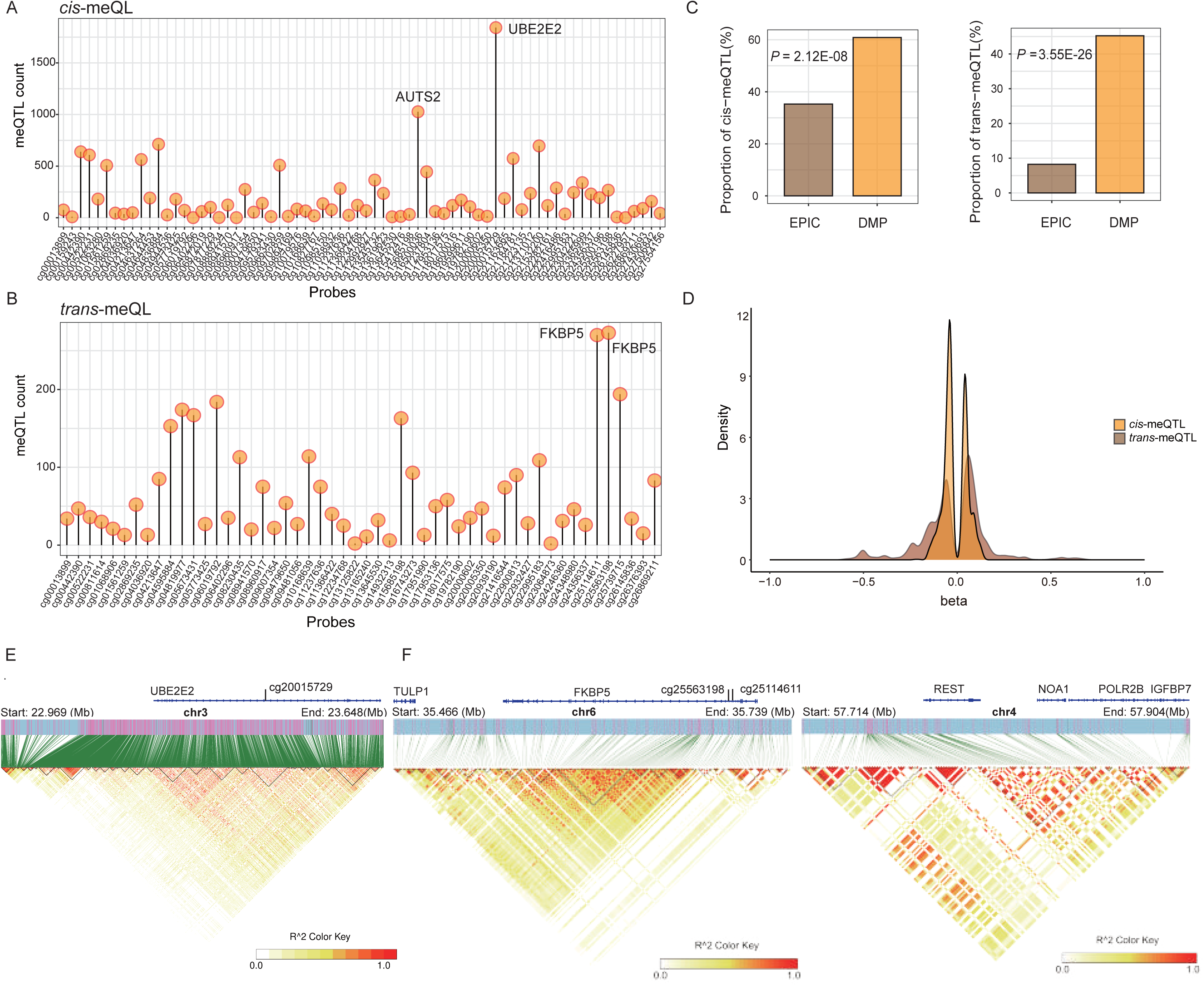
Genetic regulation of DMPs associated with CAD adverse outcomes. meQTLs associated with the 115 DMPs are listed for **(a)** cis-meQTL and **(b)** trans-meQTL. For each DMP, the number of associated meQTLs was shown. **(c)** The bar plots display the proportion of CpGs having meQTLs. **(d)** Association strength, beta, between the DMPs and the meQTLs. EPIC: all CpGs in the Infinium methylationEPIC Beadchip. DMP: 115 CpGs associated with CAD adverse outcomes. **(e)** Genomic distribution of the 1,842 cis-meQTLs associated with cg20015729, which was located on the *UBE2E2* gene. **(f)** meQTLs for two DMPs located on the promoter of the gene *FKBP5*: cg25563198 associated with 267 cis-meQTLs and 273 trans-meQTLs within chromosome 4; cg25114611 associated with 270 trans-meQTLs within chromosome 4. Linkage disequilibrium between the meQTLs, as measured by R^2, was indicated by the triangle plots.

We next assessed how these meQTLs performed in genetic association tests. We genotyped 1,551 CAD patients using Illumina GSA array, who were recruited from the three medical centers in China and partially overlap with our methylation cohort. We derived 3,448,646 high-quality single nucleotide variants (SNVs) after imputation and quality control, and performed genome-wide association studies (GWAS) with death or MACE via logistic regression (**Methods**). Among the 14,374 *cis*-meQTLs paired with the DMPs, 8,362 were genotyped and therein fewer than 5% SNPs displayed nominal association (*P* < 0.05) (see Supplementary data online, *Figure S9a-S9b*). Such weak associations would be missed by GWAS, whereas our EWAS analysis recovered them. The fact that 10s to 1000s of meQTLs regulating one single DMP, each with a weak strength, suggested that the trickling of little genetic signals had mounted to significant epigenetic outcomes, which resembled the polygenic model in complex traits. As such, the most important DMPs could be regulated by many meQTLs. Indeed, we observed that the DMP having most cis-meQTLs was cg20015729, with a remarkable 1,842 *cis*-meQTLs spanning 678.84 Kb, and closest to the gene Ubiquitin conjugating enzyme E2 E2 (*UBE2E2*). Similarly, cg16500036 was associated with over 1,000 cis-meQTLs, and closest to the gene Activator of transcription and developmental regulator (*AUTS2*). DMPs having most trans-meQTLs were cg25563198 and cg25114611, both closest to the gene FKBP Prolyl Isomerase 5 (*FKBP5*) (*Figure 4e-4f*)

Pleiotropy at the nucleotide level was frequently observed in genetic studies ^55^. We were interested in learning the SNVs that simultaneously regulated DMP methylation and gene expression, i.e., SNV with a dual role of meQTL and eQTL. Therefore, we leveraged the pleiotropic association model in SMR (**Methods**) to integrate meQTLs from Peng et al and eQTLs from eQTLGen. Briefly, 6,796 *cis*-meQTLs connected to 56 DMPs were identified as or closely located to eQTLs (*P* <1.0E-6), which regulated the expression of 242 genes. To further distinguish pleiotropy from linkage, we performed a HEIDI test against the null hypothesis that the SMR association was due to pleiotropy. As such, we revealed that 1,785 *cis*-meQTLs were indeed eQTLs (*P*_HEIDI_ >0.05), ruling out the possibility that the pleiotropic associations were caused by genotypes in linkage disequilibrium (LD). These pleiotropic meQTLs/eQTLs were linked to 29 DMPs and 71 genes, forming 80 CpG-gene pairs (see Supplementary data online, *Table S11*).

It is worth noting the SNV rs10235487 (see Supplementary data online, *Figure S9c*). As a *cis*-meQTL, it regulated cg16500036, a common feature in the CG prognostic models of death and MACE. This CpG site was located on an enhancer predicted by the ABC model to interact with *AUTS*2, therefore its increased methylation level would theoretically decrease the expression of *AUTS2*. Meanwhile, as an eQTL, this SNV negatively correlated with the expression of *AUTS2* (*P_SMR_* = 4.50E-09, *P_HEIDI_* > 0.05). Collectively, these pieces of evidence pointed to a strong connection of SNV rs10235487 to increased methylation of cg16500036 on an enhancer of *AUTS2*, which subsequently results in decreased expression of *AUTS2*, leading to poor CAD outcomes.

### Prognostic genes in acute MI and stroke

Our convergent findings presented above indicated that changes of methylation perturbed expression of the key genes involved in regulating CAD progression (see Supplementary data online, *Table S5*). To further investigate their effects, we examined expression of the 100 prognostic genes in patients of MI or stroke, two major adverse outcomes of CAD.

Bulk RNA sequencing of peripheral blood between MI patients and controls were obtained. In addition, a recent study reported single-nuclei profiling of gene expression and chromatin accessibility with spatial and time resolution, in which three zones of the MI lesion tissues were interrogated. We reanalyzed these two datasets and found that our prognostic genes displayed relatively small expression changes in MI, whether in tissue or blood, as most expression changes were within 30% (Fig. 5a-5b). This likely reflects the nature of these genes being more sentinel rather than violent players during the actual occurrence of the adverse events. A closer look showed that 27 of the 100 prognostic genes changed their expression by 30% or more in the 3 physiological zones: *myogenic*, representing non-ischemic or normal tissues; *ischemic*, the lesion site; and *fibrotic*, representing lesion sites with advanced disease progression (see Supplementary data online, *Figure S10a* and Supplementary data online, *Table S12*).

**Figure 5.**
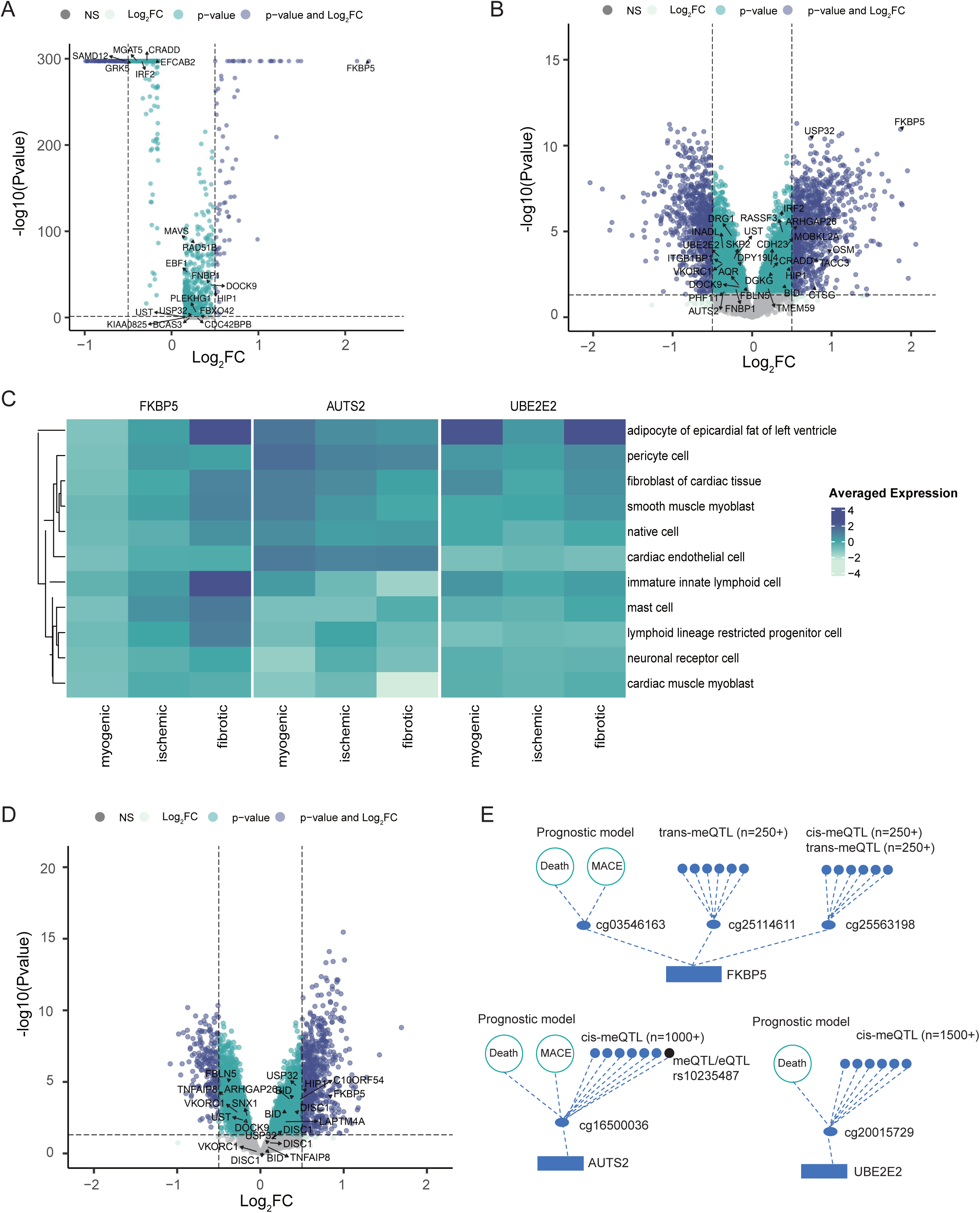
Expression of the prognostic genes in myocardial infarction (MI) and stroke. Differential gene expression in the MI patients compared to healthy controls in **(a)** MI lesion tissues and **(b)** peripheral blood. **(c)** Expression of the prognostic genes *FKBP5*, *AUTS2* and *UBE2E2* in various cell types in three locations of the MI tissues: myogenic (i.e., nonischemic zone), ischemic, and fibrotic (i.e., advanced MI zone). **(d)** Differential gene expression in the peripheral blood of stroke patients compared to the healthy controls. In all volcano plots of gene expression, prognostic genes inferred from our DNA methylation study were labeled. **(e)** A collection of evidence for the CpG sites associated with the prognostic genes *FKBP5*, *AUTS2,* and *UBE2E2*.

We also examined expression of the prognostic genes in different cell types (*Figure 5c* and Supplementary data online, *Figure S10b*). Notably, ***FKBP5***, the gene closest to the DMPs cg03546163 (a common marker in CG prognostic models for death and MACE), cg25114611 and cg25563198 (having the largest number of trans-meQTL and ABC target genes), elevated its expression in MI by four folds in nearly all cell types, and its expression levels were ever higher along the disease progression. Conversely, ***AUTS2*** nearest to cg16500036 (a common marker in the CG prognostic models for death and MACE, and serving a dual role as meQTL and eQTL), decreased its expression by 52% and 26% in immature innate lymphoid cells and smooth muscle myoblast cell in MI, and decreased gradually in immature innate lymphoid cells along the disease progression. ***UBE2E2*** nearest to cg20015729 (a marker in the CG prognostic model for death), displayed highest expression in adipocytes of the epicardial fat from the left ventricle; furthermore, we observed its expression was decreased by 20% in immature innate lymphoid cells and native cells of MI, and along the disease progression, it first decreased then elevated in several cell types, including fibroblasts of the cardiac tissue, immature innate lymphoid, and native cells, suggesting this gene may participate in cardiac repair after acute myocardial injury.

As for stroke, in a dataset where gene expression in the peripheral blood from patients of acute ischemic stroke was studied, we found 14 prognostic genes displaying differential expression as compared to the healthy controls (*Q* <0.05) (*Figure 5d*). Therein, *FKBP5* was among those having the greatest expression changes.

Therefore, there appears to be coherent evidence for the strongest signals, such as those linked to FKBP5, AUTS2, and UBE2E2. We summarized them and constructed the DMP – gene regulation models (*Figure 5e*).

## Discussion

In this study, we presented comprehensive EWAS on more than 733,000 DNA methylation sites distributed genome-wide against the survival time of adverse outcomes in 933 CAD patients. We identified 115 CpG sites whose methylation patterns were characteristic of patients with future adverse events. From a spectrum of analyses on these DMPs, we obtained three main observations.

First, we learned that 72% of the DMPs were located on enhancers and associated with genes involved in stress response, senescence, inflammation, and vessel tube regulation. An additional investigation leveraging clinical measurements revealed finer sub-categories, from which strong associations of the DMPs were observed with: (1) three inflammation indices, namely fibrinogen, SII, and PLR, all connected to platelets; (2) heart functions, namely LVEF and LVMI; and (3) HDLC. As platelets and cholesterol were essential components of thrombosis, our results suggested that early thrombo-inflammation and heart contraction functions mediated the adverse outcomes in CAD. Indeed, prognostic models based on fibrinogen, LVEF, or LVMI, each achieved AUC 0.65 or above. Interestingly, HDL is known in reverse cholesterol transport, interacts with platelets and exerts an antithrombotic function by suppressing the coagulation cascade and stimulation of clot fibrinolysis ^56^. In our analyses, DMPs were associated strongly with HDLC but little with other lipid categories; furthermore, HDLC displayed stronger power than other lipid categories in predicting adverse outcomes. As such, our study suggested that the ability to remove cholesterol, rather than its accumulation, was more relevant to the adverse outcomes. A recent study discovered that LDLC, compared to the inflammation index C-reactive protein, was less effective in predicting future cardiovascular events and death ^57^. Our study suggests that HDLC, not LDLC, may be a more relevant predictor. Therefore, our methylation study of CAD adverse outcomes may inspire new research for clinical translation.

Second, we observed a significant genetic component in the regulation of CAD adverse outcomes. Strikingly, 80% of the DMPs could be mapped to known meQTLs. In addition, important prognostic genes, which repetitively appeared as most significant in various analyses, own the largest number of meQTLs for their DMPs. For example, *UBE2E2* and *AUTS2*, each had a DMP (cg20015729 and cg16500036) associated with more than 1,000 *cis*-meQTLs. *FKBP5* had two DMPs, with one (cg25563198) associated with 267 *cis*-meQTLs and 273 *trans*-meQTLs, and the other (cg25114611) associated with 270 *trans*-meQTLs. Interestingly, our analysis showed that each of these meQTLs conferred a very weak GWAS signal; however, when combined, they collectively regulated the important DMPs. Indeed, the most important DMPs were regulated under significantly more meQTLs. Such genetic regulation of DNA methylation resembles the polygenic model in GWAS studies of many complex traits ^58^. Furthermore, we discovered that pleiotropic effects were general. In total, 55% of the meQTLs (n=6,796) that regulated the adverse outcome DMPs were in LD with known eQTLs. In fact, 15% cis-meQTLs (n=1,785) were themselves eQTLs, as assessed by the SMR and HEIDI tests. These observations suggested that a coupled genetic regulation of methylation and gene expression could be a robust mechanism.

Third, most prognostic genes displayed subtle changes during the actual adverse outcomes. This can be attributed to the nature of our study, that the biomarkers we searched mainly served as early alarms. These biomarkers represented proceeding events months to years before the adverse outcomes actually occurred. That said, however, several prognostic genes did display drastic expression changes during the adverse events and occurred repetitively as the most significant findings along various analyses. These genes include *FKBP5*, *AUTS2* and *UBE2E2*. *FKBP5* is an immunophilin protein that binds to immunosuppressive drugs. In our analysis, *FKBP5* had numerous DMPs associated with inflammation markers and heart functions. One of them, cg25114611, has been reported in acute MI ^59^, death risk ^45^, inflammatory bowel disease ^60^, Crohn’s disease ^61^, maternal BMI ^62^ and diabetes mellitus ^63^. FKBP5 expression was reported to be significantly altered in dilated cardiomyopathy after heart transplantation and suggested to serve as a prognostic marker ^64^. In our analysis of MI and stroke, *FKBP5* appeared as a most highly regulated gene and involved in pathways of cellular senescence. Its elevation in MI tissue was most drastic in innate immune lymphoid cells and adipocytes of the epicardial fat of the left ventricle. These results align with the recent finding that DNA demethylation led to increased expression of *FKBP5*, which in turn promoted NF-kB signaling in immune cells, resulting in a proinflammatory response and increased cardiovascular risk ^65^. Genetic variation in *AUTS2* was reported in blood pressure ^66^, body mass index ^66^, type 2 diabetes ^67^, and mild heart defects ^68^. *UBE2E2* was associated with type 2 diabetes ^69^, RR interval in electrocardiogram ^70^, and fat distribution ^71^. Functional analysis indicated that loss of function of *UBE2E2* in mouse primary adipose progenitor cells impaired adipocyte differentiation ^71^. Such ample connections to cardiometabolic diseases strongly support the notion that these prognostic genes played critical roles in the CAD progression.

Leveraging the DNA methylation markers and the biological insights from our analyses, we constructed succinct prognostic models for predicting death and MACE in CAD patients. To facilitate clinical application, we purposefully selected maximally 10 CpG sites; furthermore, we incorporated simple demographic information such as chronological age and sex, and a few clinical features that were relatively easy to obtain, including fibrinogen, LVEF, and HDLC. Our ensemble models achieved AUC of 0.83 for predicting death and 0.77 for predicting MACE. Furthermore, they achieved robust performance in CAD patients independently assembled from three medical centers, proving their potential for clinical translation.

There are several limitations in our study. First, most of the adverse events occurred to our CAD patients were within the first 5 years of ascertainment, therefore our study captured short-term to intermediate signals. Given a longer interrogation timespan, or a study with adverse events occurred in a longer time span, the DMPs predicting longer-term adverse events could appear. Second, the number of patients experienced adverse outcomes in the validation cohort was relatively small, i.e., there are 248 in the discovery cohort and only 41 in the validation cohort. This limited the power of replication. Indeed, 70-80% DMPs from the discovery cohort could not be replicated in the validation cohort, and therefore removed from the bioinformatic analysis and model construction. Third, although 850K EPIC array could assess CpG methylation genome-wide, many CpG sites were not probed and therefore leaves a large room for future discovery of prognostic markers.

To conclude, our study displays the value of leveraging DNA methylation of peripheral blood in predicting future adverse events in CAD patients. Further studies are warranted to investigate the roles of the methylation sites, genes, pathways, and mediating phenotypes implicated in our study for a mechanistic understanding of the CAD adverse outcomes.

## Acknowledgements

We thank the CAD patients for their willingness to consent and participate in this scientific study. We thank medical staffs of the Guangdong Provincial People’s Hospital, particularly the nurses, for their diligent and careful follow-up of the patients. We thank Dr. Sijia Wang for providing early access to the methylation QTL datasets for our research. We also thank members of the Guangdong Provincial Key Laboratory of Coronary Heart Disease Prevention and members of the Laboratory of Intelligent Computing in Biomedicine in the Greater Bay Area Institute of Precision Medicine (Guangzhou) for insightful discussions and suggestions.

## Data availability

Gene regulatory elements were obtained from ENCODE5 catalog (https://www.encodeproject.org/). Enhancer-gene predictions by the ABC models in 131 biosamples were obtained from the ENGREITZ LAB (https://www.engreitzlab.org/abc/). CpG sites associated with diseases and traits were download from EWAS Catalog (http://www.ewascatalog.org/) and EWAS Atlas (https://ngdc.cncb.ac.cn/ewas/atlas). meQTL summary statistic data were obtained from Pan-mQTL (https://www.biosino.org/panmqtl/home). eQTL summary statistics were derived from eQTLGen (https://www.eqtlgen.org/cis-eqtls.html). Single-nuclei RNA sequencing data of MI patients were available from Zenodo data archive (https://zenodo.org/record/6578047). Blood derived bulk RNA sequencing data were obtained from GEO database (<u>GSE61144</u> and <u>GSE16561</u>). Calculation of DNA methylation age of GrimAge, Hannum clock, Hovath clock, and PhenoAge were performed by DNA Methylation Age Calculator (https://dnamage.genetics.ucla.edu/home).

## Author Contributions

SZ and CP designed the study. SZ supervised and coordinated the overall study and CP supervised the data analysis. SZ, XF, BZ, XC and CL assembled the study cohort. MQ, QW, XT, QZ, XW, XC and CL consented the patients and supervised the patient follow-up. MQ, QW, XT and HL collected samples and prepared them for DNA methylation array. MQ and CP performed bioinformatic and statistical analyses and generated the figures and tables. CP and MQ drafted the manuscript. All authors contributed to result interpretation and discussions. CP, MQ and SZ critically reviewed the manuscript.

